# Countries are Clustered but Number of Tests is not Vital to Predict Global COVID-19 Confirmed Cases: A Machine Learning Approach

**DOI:** 10.1101/2020.04.24.20078238

**Authors:** Md Hasinur Rahaman Khan, Ahmed Hossain

## Abstract

COVID-19 disease is a global pandemic and it appears as pandemic for each and every nation and territory in the earth.This paper focusses on analysing the global COVID-19 data by popular machine learning techniques to know which covariates are importantly associated with the cumulative number of confirmed cases, whether the countries are clustered with respect to the covariates considered, whether the variation in the covariates are explained by any latent factor. Regression tree, cluster analysis and principal component analysis are implemented to global COVID-19 data of 133 countries obtained from the worldometer website as reported as on April 17, 2020. Our results suggest that there are four major clusters among the countries. First cluster consists of 8 countries where cumulative infected cases and deaths are highest. It is also revealed that there are two principal components. The countries which play vital role to explain the 60% variation of the total variations by the first component characterized by all variables except the rate variables include USA, Spain, Italy, France, Germany, UK, and Iran. Remaining countries contribute to explaining 20% variation of the total variations by the second component characterized by only three rate variables. We also found that the number of tests by the country variable among other variables country, number of active cases, number of deaths, number of recovered patients, number of serious cases, and number of new cases is an unimportant variable to predict cumulative number of confirmed cases. Hence, the number of tests might play vital role to individual country level who are in the primary level of virus spread but not to the global level.

## 1 Introduction

The severe acute respiratory syndrome coronavirus 2 (SARS-CoV-2) is an infectious disease which was first emerged in December 2019 in Wuhan, the capital of China’s Hubei province (Roosa et al., 2020). Since it has spreaded to nearly 213 countries and territories, has infected more than 2.3 million people by April 17, 2020, has killed approximately 155,000 people worldwide (Max Roser & Ortiz-Ospina, 2020) (also see Figure 2). As of 17 April 2020, the highest crude fatality rate was observed in Belgium (nearly 485 per million), followed by Spain (nearly 435 per million) and Italy (nearly 390 per million) (Max Roser & Ortiz-Ospina, 2020). However, the highest number of deaths took place in United States (over 38,000) followed by Italy, Spain and France. The countries who are most affected have conducted huge number of tests. As of April 17, 2020 the United States has conducted more than 3.7 million tests followed by Russia (over 1.8 million), Germany (over 1.6 million) and Italy (approximately 1.3 million). No of active cases is growing as the no. of cases are growing. As of 17 April 2020, globaly nearly 67% of the total cases are the active cases and hence 23% are the recovered (Max Roser & Ortiz-Ospina, 2020).

**Figure 1:**
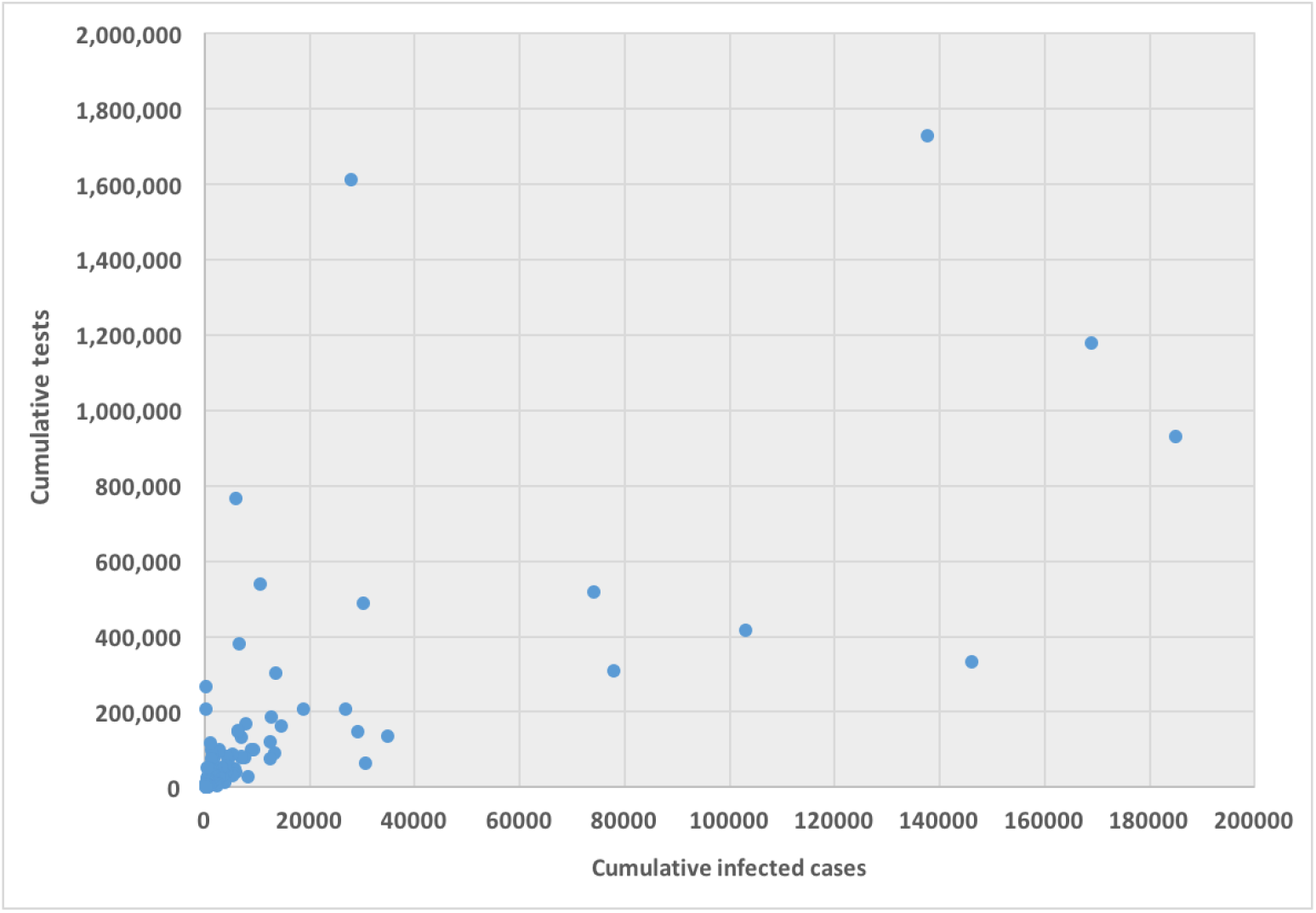
Scatter plot between cumulative tests and cumulative cases for 132 countries (except USA)

**Figure 2:**
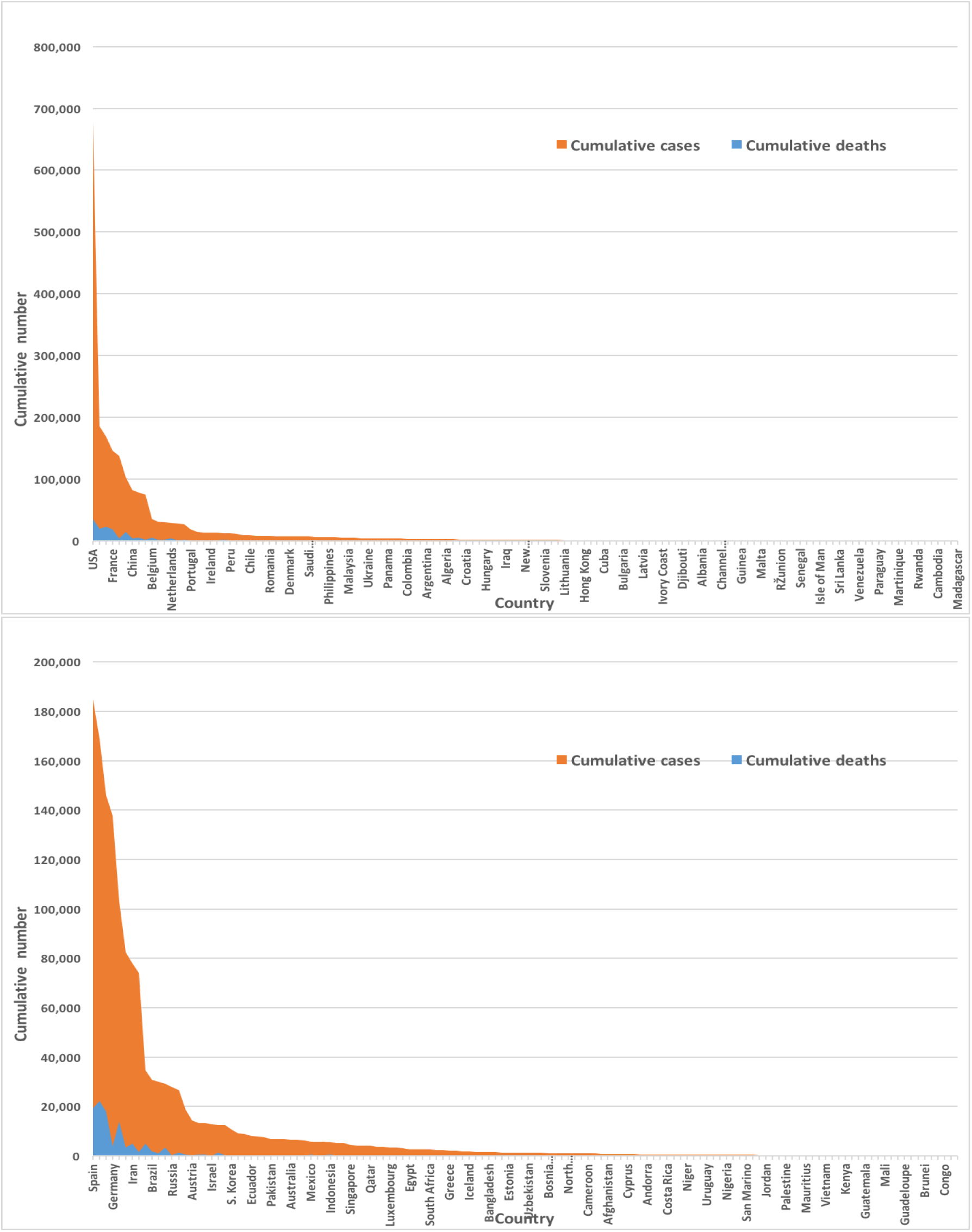
Global infected cases and deaths of COVID-19 for 133 countries (upper panel) and so without USA (lower panel) as of April 17, 2020.

Most of the affected countries have been maintaining social distancing and closing of educational institutes, offices, and markets for reducing spread in considerable rates, while these become less effective in many countries where people are commuting in crowded public transport or even living in cheek by jowl urban slums (Hui et al., 2020). Also, in many countries the public healthcare system is not sufficient and overburdened and they are in potential dangerous threat (Khan & Hossain, 2020). According to World Bank data (WB, 2020), Bangaldesh in 2015 has 0.8 hospital beds for every 1,000 people, the India has 0.7 (2011), the Pakistan has 0.6 (2012), US has 2.9 (2012) while China has 4.2 (2012) beds per 1,000 people. It is recommended that ICU practitioners, hospital administrators, governments, and policy makers must prepare for a substantial increase in critical care bed capacity, with a focus not just on infrastructure and supplies, but also on staff management (Phua et al., 2020).

Tests capability is not uniform over the countries rather haterogeneous and even within country it is heterogenous. Testing can depend on mainly country’s financial capability, laboratory capacity, and access although it is one of our most important tools for slowing down and reducing the spread and impact of the virus. Within limited capability, the low and middle income countries may have to battle their COVID-19 pandemic. Tests allow us to identify infected individuals, guiding the medical treatment that they receive. It enables the isolation of those infected and the tracing and quarantining of their contact (Hellewell et. al., 2020). As of 17 April 2020, USA administered the highest no. of tests which is approximately 3.4 million which is almost 20% of global test total, followed by Germany (over 1.7 million), Russsia (over 1.6 million) and Italy (approximately 1.2 million). Figure 1 represents the scatter plot between the cumulative cases and cumulative tests for 132 countries. USA was discarded in the graph as USA has exceptionally very high tests performed. We found correlation coefficient between these two variables for 132 countries is 0.71 that indicates strong positive correlation, while including USA it is 0.88 that indicates very very high positive correlation.

We found a number of research works where machine learning tools have used for global and local COVID-19 data. Recently, Chuanyu et al. (2020) used several machine learning tools including elastic net, random forest, and bagged flexible discriminant analysis for predicting mortality risk of COVID-19 patients. Ismail Magdon-Ismail (2020) presented a robust data-driven machine learning analysis of the COVID-19 pandemic from its early infection dynamics. “COVID-19 and artificial intelligence: protecting health-care workers and curbing the spread” (2020) discussed how artificial intelligence protecting health-care workers and curbing the spread of COVID-19. News (2020) discussed about the hunting the virus with technology, AI, and analytics. News (2020) used deep learning method for reviewing and critically appraising published and preprint reports of prediction models for COVID-19 patients. Particularly, a number of study works [Qi et al. (2020), Yan et al. (2020), Loey et al. (2020), Hu et al. (2020), IBM (2020), Fobes (2020), Healthitanalytics (2020), Corman et al. (2020), Fomsgaard & Rosenstierne (2020), Maghdid et al. (2020), Rao & Vazquez (2020), Gozes et al. (2020), Hall et al. (2020), Afshar et al. (2020), and Ghoshal & Tucker (2020)] has used machine learning including big data techniques to COVID-19 data to determine the spread of the disease, predict the risk of disease, the diagnosis of disease, number of incidence, health care faciities.

In this paper, we would like to explore whether global cumulative infected people can be predicted with the avalable data, collected as of 17 Aapril 202 from Worldometer (Max Roser & Ortiz-Ospina, 2020), on other covariates–country, number of new cases, number of active cases, numberr of deaths, number of recovered patients, number of serious cases, number of tests, deaths per million, cases per million and tests per million. If so, then we would like to know how much vital the cumulative number of tests is to predict the number of infections. We will further invesigate whether the countries are clustered on the basis of these covariates. Finaly, whether the total variations can be explained with some latent groups who are uncorrelated each other.

## 2 Methodology

The data used for the current study has been collected from the real time COVID-19 data from the Worldometer website (Max Roser & Ortiz-Ospina, 2020) until 17-th April, 2020. The Worldometer is the data repository and the free reference website which is trusted by the UK Government, Johns Hopkins CSSE etc. For the current study, we collated the information obtained on the top 133 countries with the 100 number of confirmed COVID-19 cases. For each country we collected information on total confirmed cases, new confirmed cases, total deaths, total recovered patients, total active case, total seriousely critical patients, infection rate in million, death rate in million, total tests conducted, and test rate in million. New confirmed cases are the confirmed cases reported on 17-th April. The definition of recovery and serious cases vary from country to country. According to Max Roser & Ortiz-Ospina (2020), the recovered number is not very accurate as reporting can be missing, incomplete, incorrect, based on different definitions, or dated (or a combination of all of these) for many governments, both at the local and national level, sometimes with differences between states within the same country or counties within the same state. We considered the data representing the rates such as cases, deaths and tests per million in our analysis since these are vital statistics and representing the proxy of the respective population size.

We found missing values for each variable except for the cumulative number of infected patients. There are some countries who did not provide the number of tests performed by themselves such as China, Kuwait, Oman, Cameroon, Afghanstan. Before implementing any unsupervised machine learning techniques–principal component analysis (PCA), cluster analysis and regression tree using Classification And Regression Tree (CART) method Breiman et al. (1984) by using R package caret Kuhn (2020), we imputed all missing values with the EM algorithm technique as suggested in Dray & Josse (2020).

## 3 Analysis

Figure 2 describes that most of the COVID-19 cases and deaths are from USA and counttries from Europe. We found that USA and european countries such as Germany, Russia, Italy, Spain, UK and France administered very high number of tests. The average number of tests among the available countries of 133 countries is found nearly 156,500 while USA performed the highest 3,398,140 and San Mario performed the lowest 846 tests as of April 17, 2020.

Our all variables except the country are correlated. We standardized the data and imputed the missing value through EM algorithm according to (Dray & Josse, 2020) prior to perform principal component analysis. We found principal components through orthogonal transformation by converting 133 country’s ten correlated variables into a set of values which are linearly uncorrelated variables. This exploratory data analysis is useful for making predictive models. This unsupervised machine learning technique will give the pattern of similarity in the countries and those orthogonal variables found. Figure 3 shows such pattern where the first two prinicpal components are displayed. We found that most of the variance (80%) are explained by the first two principal components.

**Figure 3:**
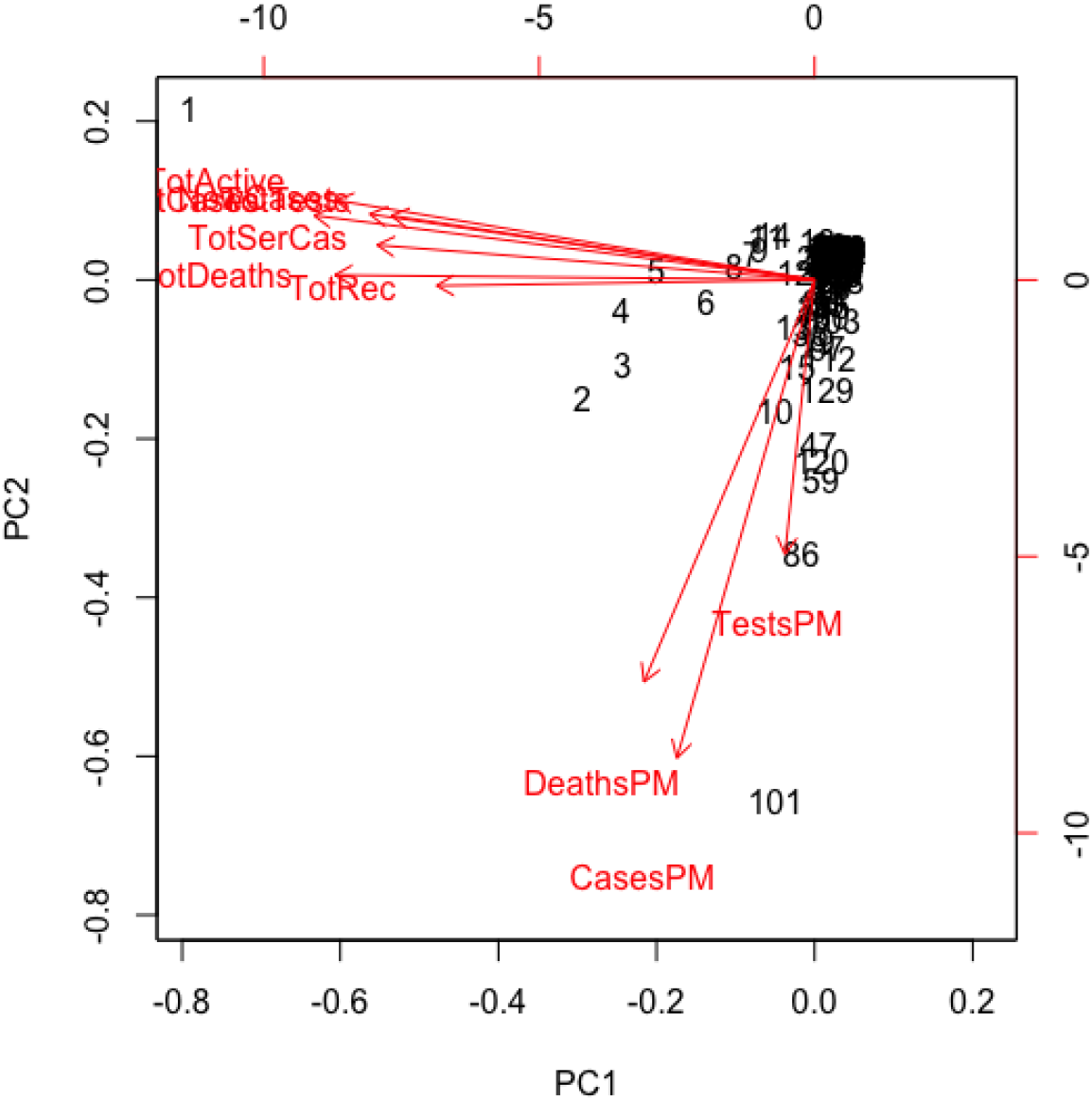
Principal component analysis results for global COVID-19 data of 133 countries.

The main results are reflected in the graph of scores of the Figure 3, where we represented the countriess in the axes formed by the first two principal components. The cloud of individual points is centered at the origin to facilitate the data analysis. The first principal component is characterized by the variables–total infected cases, deaths, active cases, recovered cases, serious cases. new cases and total tests. The countries which playing the main vital role to explain the 60% variation of total variations by the first component include USA, Spain, Italy, France, Germany, UK, and Iran. The second principal component is characterized by the remaining variables–rate of deaths, rate of infected cases and rate of tests per million. It can be noted that country’s population size as proxy of these rates playing vital role to the second principal component which explains 20% of the total variations.

We implemented cluster analysis technique to the imputed and standardized data as used in the principal component analysis. The heatmap of hirarchical cluster analysis, as shown in Figure 4, reveals that there are two clusters among the variables and four clusters among the countries. Three rate variables together– tests, cases and deaths per milllion forms one cluster while the remaining seven variables together forms the second cluster among the variables. However, we observed four mjor clusters among the countries. Table 1 shows the full list of the clusters. The first cluster contains all the countries that contributed to the first principal component’s variation in PCA analysis along with China. The PCA also suggests to valid this clustering because the heatmap in Figure 4 reveals that these countries are clustered based on the maximum variation directed by the all seven variables. The second cluster contains 43 countries which are clustered according to all variables except for the test and case rates per million. The third cluster consists of 14 countries that are clustered based on all variables other than death rate in per million. These countries have very lower deaths. The final cluster is consisted of the highest number of 68 countries which are clustered mainly based on the test and case rates variable along with other variabless used in this study.

**Table 1:** Cluster-wise country lists for 133 countries

| Cluster | Country |
| --- | --- |
| Cluster 1<br>(n=8) | USA, Spain, Italy, France, Germany, UK, China, Iran |
| Cluster 2<br>(n=43) | Afghanistan, Australia, Austria, Belarus, Brazil, Brunei, Burkina Faso, Cameroon, Canada, Congo, Cyprus, Czechia, Denmark, Diamond Princess, DRC, Estonia, Finland, Guadeloupe, Guinea, Hong Kong, Israel, Ivory Coast, Kuwait, Latvia, Lithuania, Madagascar, Mali, Martinique, Netherlands, New Zealand, Norway, Oman, Portugal, Qatar, Russia, Runion, S. Korea, Senegal, Singapore, Slovenia, Sweden, Turkey, Venezuela |
| Cluster 3<br>(n=14) | Andorra, Bahrain, Belgium, Channel Islands, Faeroe Islands, Gibraltar, Iceland, Ireland, Isle of Man, Luxembourg, Malta, San Marino, Switzerland, UAE |
| Cluster 4<br>(n=68) | Albania, Algeria, Argentina, Armenia, Azerbaijan, Bangladesh, Bolivia, Bosnia and Herzegovina, Bulgaria, Cambodia, Chile, Colombia, Costa Rica, Croatia, Cuba, Djibouti, Dominican Republic, Ecuador, Egypt, El Salvador, Georgia, Ghana, Greece, Guatemala, Honduras, Hungary, India, Indonesia, Iraq, Jamaica, Japan, Jordan, Kazakhstan, Kenya, Kyrgyzstan, Lebanon, Malaysia, Mauritius, Mayotte, Mexico, Moldova, Montenegro, Morocco, Niger, Nigeria, North Macedonia, Pakistan, Palestine, Panama, Paraguay, Peru, Philippines, Poland, Romania, Rwanda, Saudi Arabia, Serbia, Slovakia, South Africa, Sri Lanka, Taiwan, Thailand, Trinidad and Tobago, Tunisia, Ukraine, Uruguay, Uzbekistan, Vietnam |

**Figure 4:**
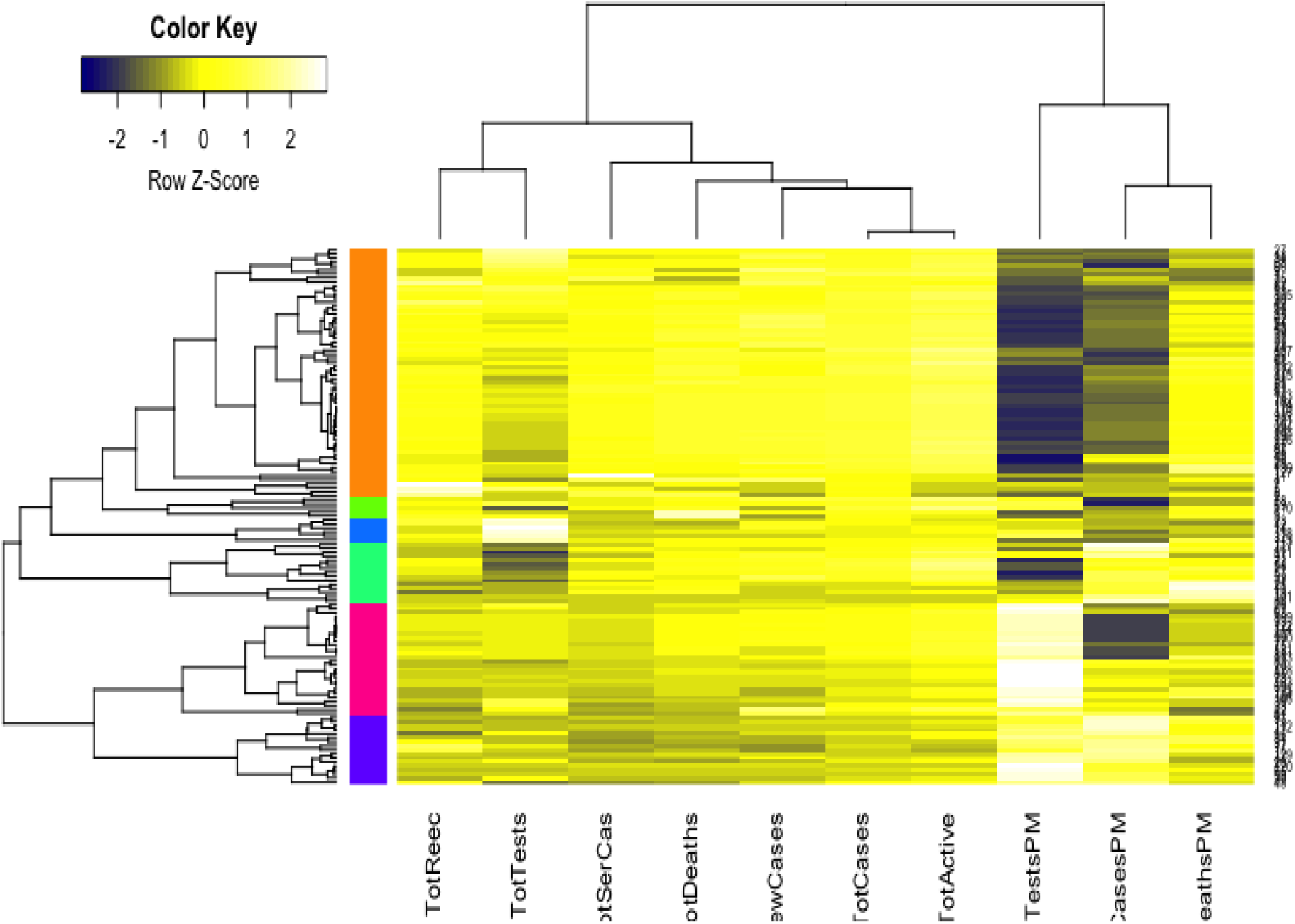
Cluster analysis results for global COVID-19 data of 133 countries.

We implemented the regression tree using CART to predict the cumulative number of infected people. Main purpose of implementing regression tree is to see whether the global cumulative number of infected people can be predicted very well with the ten variables under study. Results are presented in Table 2 that shows the weights including their percentage of importance for all ten variables. It reveals from the results that country and cumulative active cases appear as the most important variables to predict the cumulative number of infected people, followed by cumulative deaths, cumulative recovered cases, new case and cumulative serious cases. However, most strikingly we found that the cumulative tests appears as one of the most unimportant variablesto predict the cumulative number of infections.

**Table 2:**
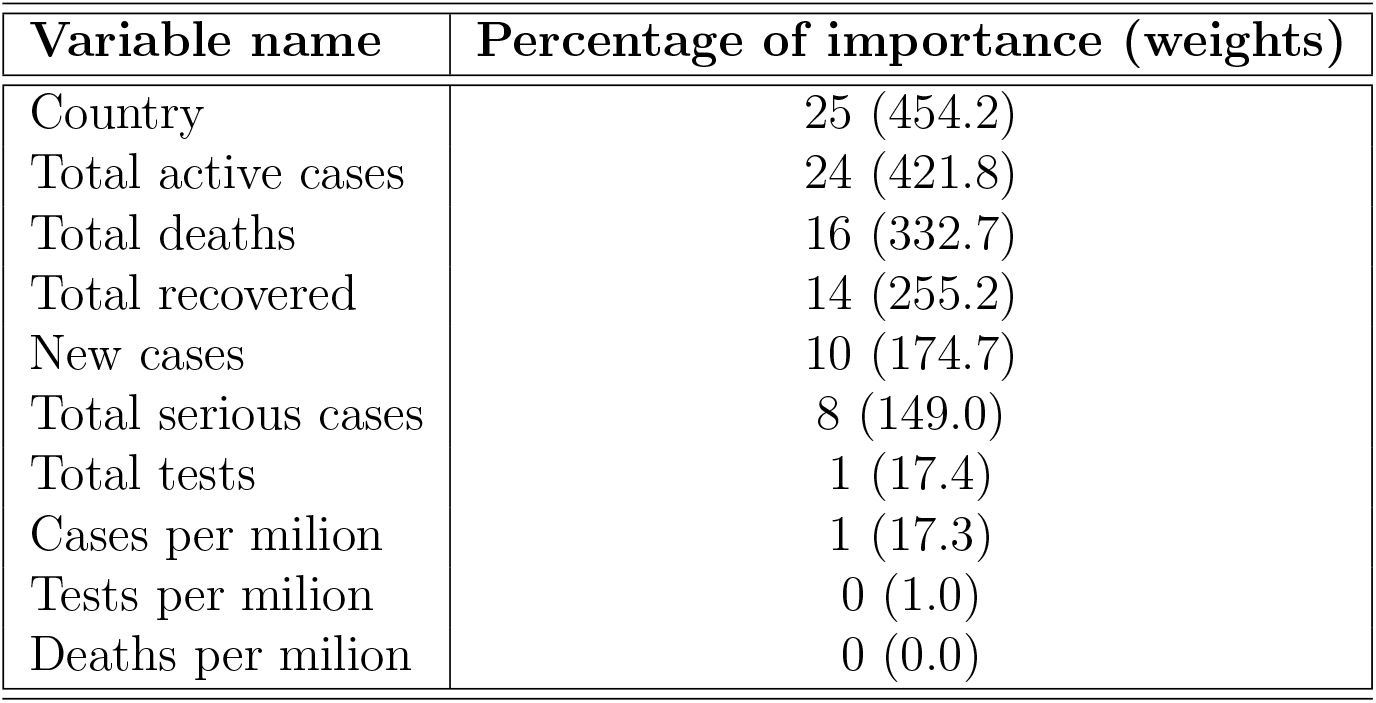
Importance of variables by regression tree

| Variable name | Percentage of importance (weights) |
| --- | --- |
| Country | 25 (454.2) |
| Total active cases | 24 (421.8) |
| Total deaths | 16 (332.7) |
| Total recovered | 14 (255.2) |
| New cases | 10 (174.7) |
| Total serious cases | 8 (149.0) |
| Total tests | 1 (17.4) |
| Cases per milion | 1 (17.3) |
| Tests per milion | 0 (1.0) |
| Deaths per milion | 0 (0.0) |

## 4 Discussions and Conclusions

In this paper, we demonstrated how to implement the basic machine learning techniques– principal component, cluster analysis and regression tree to analyse global COVID-19 data that was extracted from the Worldometer website (Max Roser & Ortiz-Ospina, 2020) and reported as of April 17, 2020. We considered 10 variables for each of 133 countries. We found from the PCA analysis that there are two latent variables that are characterized by the 10 variables we considered. The first principal component explains 60% variation of the total variations, while this is characterized mainly by 7 variables. These are the total infected cases, deaths, active cases, recovered cases, serious cases. new cases and total tests. The source of the majority of total variations is collectively all variables but the rate variables. Remaining three variables– case, death and test rates measured in per million characterize the second principal component that is due for the 20% variation of the total variations. The latent factor behind this appears to be the country’s population size as all these three variables are the proximates to population size. Neither populations of 133 countries are uniform nor the population density. We belive that country’s population size or indirectly the associated population density is responsible for the 20% variation of the total variations.

The cluster analysis found four major clusters among the countries but two clusters among the 11 variables. It reveals from the analysis that the countries are clustered based on the variation among the variables. We found that the 8 countries which having the highest number of cases form a cluster, while 43 countries form another cluster based on mainly all the variables but the case rate and test rate. The eight countries are USA, Spain, Italy, France, Germany, UK, China and Iran who are homogeneous interms of cumulative cases, deaths, active cases and tests. Most of them were/are the epicenter of the pandemic. However, we found that 14 countries, who have very low rate of deaths, form one cluster and 68 countries who have higher test and case rates along with significant effect of other eight variables form the fourth cluster. Countries having low death rates includes Bahrain, Belgium, Channel Islands, Faeroe Islands, Gibraltar, Iceland, Ireland, Isle of Man, Luxembourg, Malta, San Marino, Switzerland, UAE.

We found from the regression tree results that country, total active cases, total deaths, total recovered cases, new cases and total serious cases are very important variables to predict the cumulative number of cases but the number of tests including three rate variables is not the important variable. As stated, global data analysis indicates that the cumulative number of tests is not significant to predict cumulative cases but it is quite important to consider a specific country is in what situation or context. Besides, the policies on testing differs from country to country, region to region or even city to city. It mainly depends on what stage that country or community has reached in the pandemic curve and at the same time the level of preparedness in the specific context like number of lab facility, lab staff, sample collection strategy etc. When resources are limited and when the healthcare system is overloaded the widespread testing as suggested by WHO may not be implemented. This is a reality to many low and middle income countries in the list of 133 countries in our study. Aparently, number of tests is very important for many countries to limit the spread in early stage or even in any stage of spreading by identifying cases and isolating them and their contacts. However, global COVID-19 data analysis results revealing that cumulative tests is not at all any important determinant to predict the cumulative number of tests for the country.

The world grapples with the containment of the COVID-19 outbreak and countries are trying to reduce virus spread by performing tests for detecting and then isolating the infected people and quaranting the susceptible people. Besides, cntinueing the lockdown and social distancing is expected to help in reducing the spread in considerable amount. However, this paper found that the countries are clustered with respect to underlying effects of the covariates although the countries are fighting independently against this virus war. Similarly, variables related to rates is together a cluster while other variables together is another cluster of variables. Most strikingly, we found that the cumulative tests appeared as an unimportant variable to predict the cumulative infected people.

## Data Availability

The working data set used for this study has been submitted to the journal as additional supporting file.

## Author’s Contributions

MHRK carried out the statistical analysis and contributed to draft the manuscript. AH arranged the datasets and contributed to finalize the manuscript.

